# Eco-epidemiological assessment of the COVID-19 epidemic in China, January-February 2020

**DOI:** 10.1101/2020.03.29.20046565

**Authors:** Peter Byass

## Abstract

**Background:** The outbreak of COVID-19 in China in early 2020 provides a rich data source for exploring the ecological determinants of this new infection, which may be of relevance elsewhere.

**Objectives:** Assessing the spread of the COVID-19 across China, in relation to associations between cases and ecological factors including population density, temperature, solar radiation and precipitation.

**Methods:** Open-access COVID-19 case data include 18,069 geo-located cases in China during January and February 2020, which were mapped onto a 0.25° latitude/longitude grid together with population and weather data (temperature, solar radiation and precipitation). Of 15,539 grid cells, 559 (3.6%) contained at least one case, and these were used to construct a Poisson regression model of cell-weeks. Weather parameters were taken for the preceding week given the established 5-7 day incubation period for COVID-19. The dependent variable in the Poisson model was incident cases per cell-week and exposure was cell population, allowing for clustering of cells over weeks, to give incidence rate ratios.

**Results:** The overall COVID-19 incidence rate in cells with confirmed cases was 0.12 per 1,000. There was a single case in 113/559 (20.2%) of cells, while two grid cells recorded over 1,000 cases. Weekly means of maximum daily temperature varied from −28.0 to 30.1 °C, minimum daily temperature from −42.4 to 23.0 °C, maximum solar radiation from 0.04 to 2.74 MJm^−2^ and total precipitation from 0 to 72.6 mm. Adjusted incidence rate ratios suggested brighter, warmer and drier conditions were associated with lower incidence.

**Conclusion:** Though not demonstrating cause and effect, there were appreciable associations between weather and COVID-19 incidence during the epidemic in China. This does not mean the pandemic will go away with summer weather but demonstrates the importance of using weather conditions in understanding and forecasting the spread of COVID-19.

## Background

In infectious outbreak situations, much epidemiological effort rightly goes into case-finding and follow-up in order to track epidemics. Population-based analyses of ecological factors are however also important, not least to inform models and prognostics for future spread of the same infection elsewhere. Where a new disease is involved, such as COVID-19, it is particularly important to chart an unknown infectious agent’s interactions with environments in which transmission has occurred.

The large-scale outbreak of COVID-19 in China at the start of 2020, by now largely contained, presents an important opportunity to carry out an eco-epidemiological assessment which may be relevant for understanding patterns of transmission relevant for the ensuing pandemic. Unprecedented open-access data at the individual case level for the outbreak in China, including geo-location data, plus the availability of detailed remote-sensed and global-gridded ecological data, make this possible.

Many well-established pathogens follow well-known seasonally and ecologically determined patterns of activity. [1] Indeed, the now largely out-dated concept of “tropical medicine” was largely predicated around pathogens and vectors predominantly encountered in tropical regions during the colonial era, with some cases in travellers manifesting elsewhere. [2] Obviously little is yet known about seasonal and ecological patterns for the new SARS-CoV-2 coronavirus causing the current COVID-19 pandemic, although it is already clear that this pathogen has capacity for wide and rapid geographic spread. However, other coronaviruses, such as Middle East Respiratory Syndrome Coronavirus (MERS-CoV) have been shown to follow established seasonal patterns. [3] Human coronavirus infections have been found to be more common in winter in Norway, [4] and in Israel in summer. [5] Thus other coronaviruses show various seasonal transmission patterns.

The original COVID-19 epicentre in Wuhan City reported approximately 3 times as many cases as the whole of the rest of China, with peak incidence during January 2020. [6] This overwhelming number of cases in a single location was not included in analyses here since it did not contribute to geographic variation, and occurred earlier than the generalised epidemic in China.

The objective of this eco-epidemiological assessment was to use data on COVID-19 incident cases throughout China during January and February 2020 (excluding the original epidemic focus of Wuhan city) and to relate them to week of confirmation, population and meteorological data. This represents a kind of “natural experiment” in terms of how secondary epidemic foci occurred in diverse locations around China, before the national epidemic was more or less brought under control by the end of February, and how location-specific incidence rates varied.

## Methods

This assessment is based on the open-access COVID-19 incident case data maintained by the Open COVID-19 Data Curation Group. [7] A total of 18,069 geo-located COVID-19 incident confirmed cases during January and February 2020 were extracted for the whole of China, excluding Wuhan City. No case confirmations were reported for the first two weeks of January. All cases were mapped by week of confirmation onto a 0.25° latitude/longitude grid (approximately 25 × 25 km squares) for the whole of China, which included 15,539 cells. This grid was also filled with population data from the NASA Socioeconomic Data and Applications Center. [8] Gridded weather data for the whole of China during January and February 2020 were sourced from the Copernicus Climate Change Service ERA-5T model, specifically temperature at 2m, total precipitation and total sky direct solar radiation at surface. [9] Weekly averages/totals of daily weather data for each geographic cell were calculated and mapped on the geographic grid. These data were used as the basis for generating maps of cases and ecological factors.

There were 559/15,539 (3.6%) grid cells which contained at least one reported case at some point during January and February (excluding Wuhan City), and these were used to construct a Poisson regression model (Stata 12) in which the unit of observation was cell-week, over the period during which cases occurred anywhere in the country (weeks 3 to 9 of 2020, total 3,913 units of observation). Weather parameters during the preceding week were included in the model on the basis of the established 5-7 day incubation period for COVID-19, since this was likely to reflect weather at the time of disease transmission. [10] The dependent variable in the Poisson model was the number of incident cases in the cell-week and exposure was the population in the cell, allowing for clustering by the grid cell identifier over weeks, with incidence rate ratio as the outcome. In the absence of established hypotheses on relationships between ecological factors and SARS-CoV-2 transmission, tertiles of quantitative variables were constructed as independent variables to avoid erroneously imposing linear assumptions. Grid cells which had no incident cases reported throughout January and February were excluded, in the absence of any evidence of the possibility of transmission in the cell.

## Results

A total of 18,069 confirmed cases were reported, contained in 559 0.25° grid cells, which corresponded to an overall population of 151.2 million, around 10% of the total Chinese population. This amounted to an overall COVID-19 incidence rate in cells with confirmed cases of 0.12 per 1,000 population. In 113/559 (20.2%) of grid cells with cases, there was only one case during the whole period, while two grid cells recorded over 1,000 cases.

Figure 1 shows [a] the geographic distribution of cases, [b] population density for the whole country, and, for January-February 2020, [c] average of daily maximum temperature at 2m, [d] average of daily minimum temperature at 2m, [e] total precipitation and [f] average of daily total sky direct solar radiation at the surface. The Supplementary Material contains similar maps for temperature, precipitation and solar radiation on a weekly basis. Weekly means of maximum daily temperature varied from −28.0 to 30.1 °C, minimum daily temperature from −42.4 to 23.0 °C, maximum solar radiation from 0.04 to 2.74 MJm^−2^ and total precipitation from 0 to 72.6 mm.

**Figure 1:**
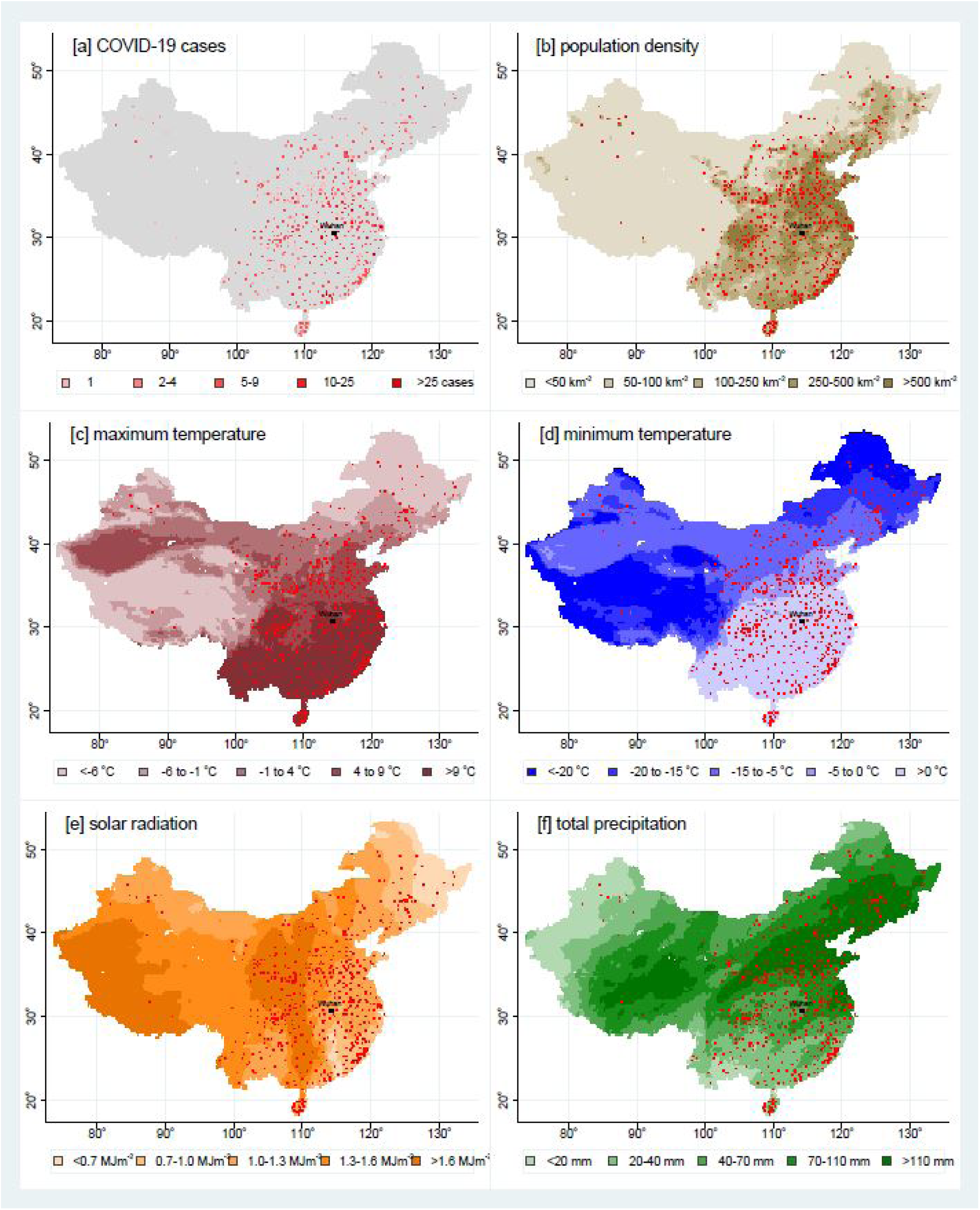
Maps of China, based on 15,539 0.25° grid cells, showing, for January-February 2020, [a] cell-densities of COVID-19 cases, [b] population density, [c] maximum temperature, [d] minimum temperature, [e] solar radiation and [f] precipitation. Maps [b] – [f] show the 559 cells having at least one case in red.

Table 1 summarises the data included in building a Poisson regression model. Because of concerns about collinearity between maximum and minimum temperatures and solar radiation for constructing a multivariable regression model, average daily temperature was calculated as the average of maximum and minimum daily temperatures, as a single measure of temperature. Then, to avoid imposing linear relationships between temperature, solar radiation and COVID-19 incidence, which appeared improbable from the bivariable results, a tertiles of tertiles approach was used, in which tertiles of average temperature were further broken down into tertiles of solar radiation, as shown in Figure 2. The nine categories derived in this way were then used as a categorical independent variable in the regression model, rather than having separate competing variables for temperature and solar radiation.

**Table 1:** Details of 3,913 0.25° grid cell-weeks of observation for COVID-19 incident cases in China (excluding Wuhan City) during January-February 2020, covering a total of 18,069 cases among a population of 151.2 million. All grid cells in which at least one case was observed during the overall period of observation are included. Bivariate incidence rate ratios and their 95% confidence intervals were calculated using a Poisson regression model with the weekly number of cases as the dependent variable, the grid cell population as the exposure variable and the previous week’s weather data as independent variables.

| parameter | level | units | range | median | bivariate incidence rate ratio | 95% confidence interval |
| --- | --- | --- | --- | --- | --- | --- |
| latitude |  | degrees | 18.25 to 49.75 | 32.25 |  |  |
| longitude |  | degrees | 82.00 to 132.25 | 113.25 |  |  |
| population | per grid cell | n | 1,169 to 2.56 million | 187,616 |  |  |
| COVID-19 cases | per grid cell | n | 1 to 1,262 | 6 |  |  |
| week in 2020 | 3 |  |  |  | 1 (ref) | – |
|  | 4 |  |  |  | 27.5 | 22.7 – 33.4 |
|  | 5 |  |  |  | 84.9 | 70.2 – 102.7 |
|  | 6 |  |  |  | 38.9 | 32.1 – 47.0 |
|  | 7 |  |  |  | 11.8 | 9.7 – 14.4 |
|  | 8 |  |  |  | 1.80 | 1.4 – 2.3 |
|  | 9 |  |  |  | 1.38 | 1.1 – 1.8 |
| population density | 1<sup>st</sup> quintile | km<sup>-2</sup> | 2 to 109 | 57 | 1 (ref) | – |
|  | 2<sup>nd</sup> quintile | km<sup>-2</sup> | 110 to 198 | 155 | 0.75 | 0.69 – 0.81 |
|  | 3<sup>rd</sup> quintile | km<sup>-2</sup> | 199 to 392 | 275 | 1.01 | 0.94 – 1.08 |
|  | 4<sup>th</sup> quintile | km<sup>-2</sup> | 393 to 631 | 489 | 0.67 | 0.63 – 0.72 |
|  | 5<sup>th</sup> quintile | km<sup>-2</sup> | 632 to 3,626 | 788 | 0.20 | 0.18 – 0.21 |
| preceding week mean of daily maximum temperature | 1<sup>st</sup> tertile | °C | -18.1 to 5.2 | -0.3 | 1 (ref) | – |
|  | 2<sup>nd</sup> tertile | °C | 5.2 to 11.7 | 8.5 | 2.11 | 2.02 – 2.20 |
|  | 3<sup>rd</sup> tertile | °C | 11.7 to 29.2 | 16.1 | 1.00 | 0.96 – 1.05 |
| preceding week mean of daily minimum temperature | 1<sup>st</sup> tertile | °C | -31.2 to -3.1 | -11.3 | 1 (ref) | – |
|  | 2<sup>nd</sup> tertile | °C | -3.1 to 4.5 | 1.4 | 3.27 | 3.11 – 3.42 |
|  | 3<sup>rd</sup> tertile | °C | 4.5 to 15.9 | 8.3 | 1.13 | 1.07 – 1.19 |
| preceding week mean of daily average temperature | 1<sup>st</sup> tertile | °C | -22.4 to 1.2 | -5.7 | 1 (ref) | – |
|  | 2<sup>nd</sup> tertile | °C | 1.2 to 8.1 | 4.8 | 3.58 | 3.42 – 3.76 |
|  | 3<sup>rd</sup> tertile | °C | 8.1 to 19.4 | 12.0 | 1.11 | 1.05 – 1.17 |
| preceding week mean of daily max solar surface radiation | 1<sup>st</sup> tertile | MJm<sup>-2</sup> | 0.07 to 0.82 | 0.59 | 1 (ref) | – |
|  | 2<sup>nd</sup> tertile | MJm<sup>-2</sup> | 0.59 to 1.33 | 1.07 | 2.57 | 2.48 – 2.67 |
|  | 3<sup>rd</sup> tertile | MJm<sup>-2</sup> | 1.33 to 2.44 | 1.64 | 0.72 | 0.69 – 0.76 |
| preceding week total precipitation | 1<sup>st</sup> tertile | mm | 0 to 4.1 | 1.8 | 1 (ref) | – |
|  | 2<sup>nd</sup> tertile | mm | 4.1 to 11.4 | 7.2 | 2.57 | 2.44 – 2.70 |
|  | 3<sup>rd</sup> tertile | mm | 11.4 to 55.9 | 17.2 | 4.71 | 4.50 – 4.94 |

**Figure 2:**
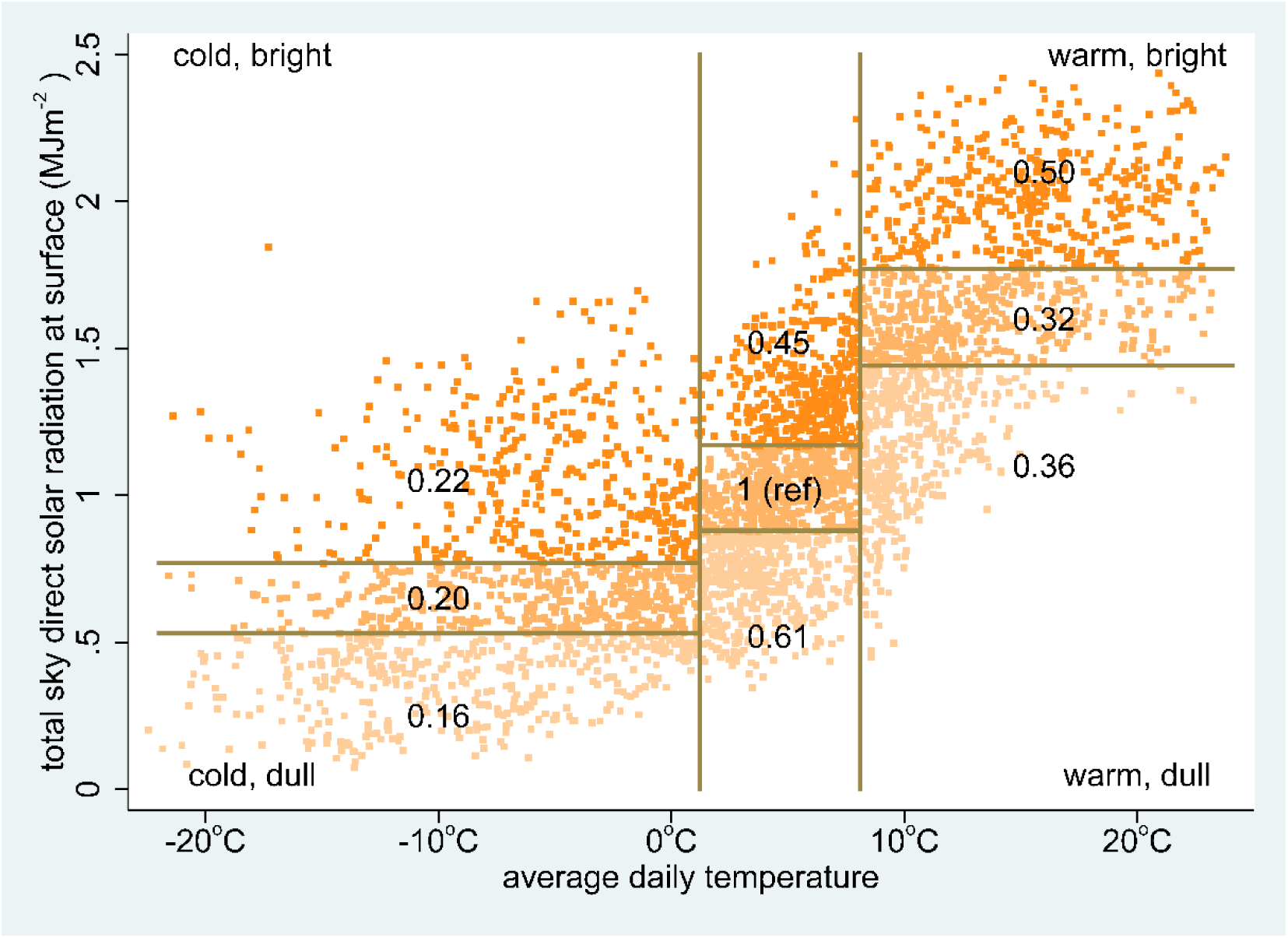
Weekly means of average daily temperature and solar radiation for 3,913 0.25° grid cell-weeks (shown as dots) of observation for COVID-19 incident cases in China (excluding Wuhan City) during January-February 2020, covering a total of 18,069 cases among a population of 151.2 million. Temperature is divided into tertiles, with each tertile then divided into tertiles of solar radiation. Numbers in each sector represent COVID-19 incidence rate ratios (adjusted for adjusted for week, population density and precipitation).

Figure 3 shows adjusted incidence rate ratios and 95% confidence intervals for the overall regression model. Adjusted incidence rate ratios (adjusted for week, population density and precipitation) for the composite temperature-radiation variable from the regression model are also shown in Figure 2.

**Figure 3:**
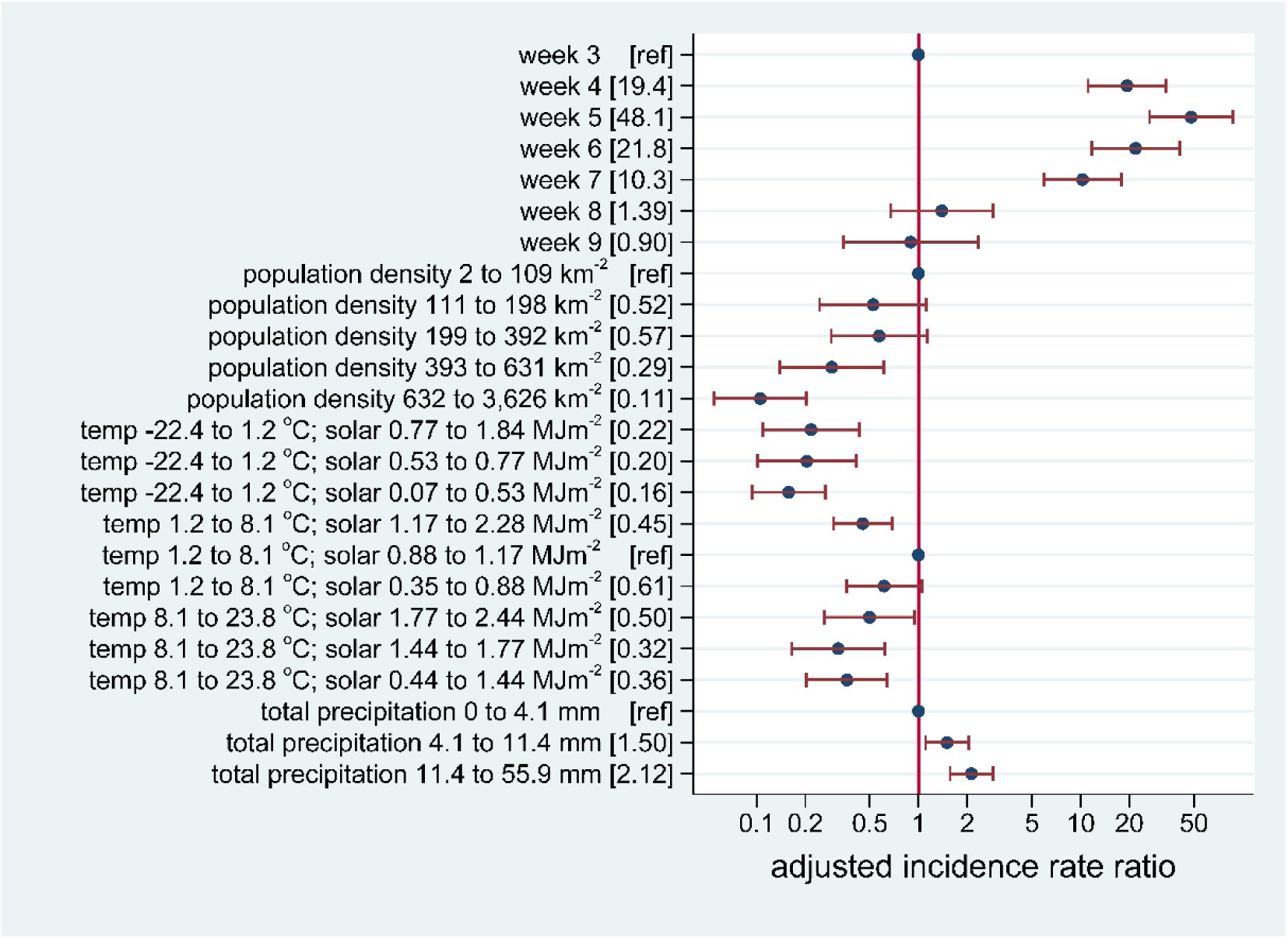
Adjusted incidence rate ratios with 95% confidence intervals in relation to weeks, population density, temperature, solar radiation and precipitation for a total of 18,069 COVID-19 cases in 559 0.25°grid cells, corresponding to a population of 151.2 million, in China (excluding Wuhan City) during January-February 2020.

Most of the adjusted incidence rate ratios were significantly different from 1, evident in Figure 3 from the lack of overlap of 95% confidence intervals with the vertical line at an adjusted incidence rate ratio of 1.

The weather parameters for Wuhan City during the large epidemic peak there in January were average daily temperature 2.0 °C, solar radiation 0.93 MJm^-2^ and precipitation 32 mm. This corresponds to the highest incidence rate ratio categories in the assessment for the rest of China shown in Figure 3.

## Discussion

These analyses clearly show variations in COVID-19 case incidence rates in China which are associated with weather during the week preceding case confirmation. An observational study of this kind cannot formally attribute cause and effect, and there remain uncertainties which have to be considered as unknown unknowns. Nevertheless, this assessment of the effects of weather on COVID-19 transmission in China suggests variations of a sufficient magnitude to have important possible consequences for understanding the current COVID-19 pandemic in other settings. This assessment also illustrates the value of the detailed open-access data available on COVID-19 cases and ecological parameters.

While the open-access individual case data is a hugely valuable resource, it is not able to tell the full story of the circumstances of each case. The nature of COVID-19 transmission is such that many cases will have no idea exactly where, when or how they acquired their infection, and so the location data for cases are inevitably more reflective of illness rather than infection. During the period of COVID-19 spread around China, following the initial epicentre epidemic in Wuhan City, the Chinese authorities implemented stringent infection reduction measures in many locations, the exact nature and chronology of which are not documented. The somewhat counterintuitive relationship seen in this assessment between population density and COVID-19 incidence very likely reflects the effectiveness of infection control measures targeted at densely populated urban areas. However, records of cases in 559 locations outside Wuhan City, as shown in Figure 1[a], show that there was widespread nationwide transmission, even though only a single case was recorded in 20% of locations with cases, which probably speaks to the effectiveness of control measures in many places.

Despite possible weaknesses around the case data, one of the strengths of this assessment is that all the other data were sourced from global data models that are totally independent of the COVID-19 data from China. Additionally, since China is a very large and geographically varied country, assessment of COVD-19 incidence was made over a very wide range of weather, as evident from Figure 1[c-f].

Among the unknown unknowns, the effect of weather on human behaviour, and thus on behavioural risks for acquiring COVID-19, may also be important. The incidence of infections in China was markedly lower at very low temperatures, which might be related to characteristics of the virus, but equally may reflect reduced social interactions when it is very cold outside. Conversely brighter, drier weather may stimulate levels of social interaction, and thereby possibly counteract direct effects of heat and light on viruses. The complex relationship between temperature and solar radiation, as shown in Figure 2, is important, because the effects of temperature and light, particularly in the ultra-violet spectrum, have been shown to be associated with seasonal viral activity in other contexts. [11] In this assessment, the independent association of precipitation with COVID-19 incidence rates was also appreciable. A recent meteorological analysis showed very similar weather patterns across a number of COVID-19 hotspots, including Wuhan City, in a corridor 30° to 50° North in early 2020. [12] This is congruent with findings here that weather conditions in Wuhan City during January corresponded to the highest risk categories assessed across the rest of China.

## Conclusion

While this assessment showed appreciable associations between COVID-19 incidence rates and weather patterns in China during January-February, this does not amount to establishing a clear cause and effect relationship. Neither does it justify any generalisations to the effect that the COVID-19 pandemic will simply go away given some nice summer weather. However, the size of the associations between weather and incidence in China are very much of public health interest in understanding the continuing spread of the SARS-CoV-2 virus around the world, in different climate zones. The possibility that transmission is reduced during periods of brighter, warmer, drier weather is important. Further assessments of this kind in other locations and seasons are needed to build a full picture, but meteorological data should be considered for inclusion in overall models of COVID-19 epidemiology and spread of the pandemic.

## Data Availability

All the data used are already in the public domain and available as specified in the manuscript

## Ethical approval

No ethical approval was required for this study, which relied solely on open-access public data.

